# Rumination as a cognitive vulnerability factor in perinatal bereavement: evidence from the CARING study

**DOI:** 10.64898/2026.06.16.26355798

**Authors:** Claudia Ravaldi, Laura Mosconi, Giorgia Raduzzi, Cristina Olmi, Ilaria Neri, Elena Cocchi, Alfredo Vannacci

## Abstract

**Purpose:** Perinatal loss is associated with a high risk of persistent psychological distress, including prolonged grief, depression, anxiety, and post-traumatic stress symptoms. Cognitive processes such as rumination may play a crucial role in maintaining and amplifying distress following loss, yet their specific contribution in perinatal bereavement remains underexplored.

**Methods:** The CARING (Cognitive Analysis and Rumination INvestigation in perinatal Grief) study employed a cross-sectional design involving 298 parents who experienced perinatal loss within the previous five years. Participants completed an anonymous online survey including measures of depressive rumination (Ruminative Response Scale, RRS), angry rumination (Anger Rumination Scale, ARS), perinatal grief (Perinatal Grief Scale, PGS), general psychopathology (SCL-90), and post-traumatic stress symptoms (NSESSS). Non-parametric analyses were conducted to examine associations between rumination patterns and psychological outcomes.

**Results:** Higher levels of rumination were significantly associated with greater perinatal grief, depressive and anxiety symptoms, and post-traumatic stress. Depressive rumination showed consistently stronger associations with all outcomes compared to angry rumination. Participants presenting both depressive and angry rumination exhibited the highest levels of grief intensity, psychological distress, and PTSD symptoms, suggesting a graded relationship between rumination patterns and severity of distress. Rumination levels were not significantly associated with gestational age at loss or with having received psychological support.

**Conclusions:** Rumination, particularly in its depressive form, appears to function as a transdiagnostic cognitive vulnerability factor in perinatal bereavement. These findings highlight rumination as a potential target for early screening and tailored psychological interventions aimed at reducing long-term distress following perinatal loss.

## 1 Introduction

Perinatal loss represents a profound psychological event that occurs at a time characterized by intense emotional, relational, and identity investment in the expected child. Loss during pregnancy or shortly after birth disrupts parental expectations and future-oriented representations, placing parents at heightened risk for persistent psychological distress. A growing body of literature has documented the significant mental health impact of perinatal loss, with consistently elevated rates of depression, anxiety, complicated grief, and post-traumatic stress symptoms, particularly among mothers (Kersting and Wagner 2012). A recent systematic review and meta-analysis found that perinatal loss is associated with elevated levels of depressive and anxiety symptoms compared to controls and highlighted the substantial psychological burden experienced by bereaved parents across multiple studies and settings (Herbert et al. 2022). These findings underscore the importance of examining cognitive and emotional processes that may influence grief outcomes.

Beyond the emotional impact of the loss itself, cognitive and metacognitive processes may critically shape the course of psychological adaptation. Among these, rumination (defined as repetitive, involuntary, automatic, and abstract thinking focused on negative emotional states and their causes or consequences) has been identified as a key vulnerability factor across multiple forms of psychopathology (Nolen-Hoeksema 1991; Watkins 2008). Rumination has been shown to contribute to the onset, maintenance, and exacerbation of depressive symptoms, anxiety, and post-traumatic stress, by reinforcing negative self-appraisals, impairing problem-solving, and sustaining emotional arousal.

Rumination is not a unitary construct. Depressive rumination typically involves self-focused, past-oriented thoughts centered on loss, failure, and perceived personal inadequacy, whereas angry rumination focuses on perceived injustice, blame, and resentment related to the event (Sukhodolsky et al. 2001). While both forms have been associated with psychological distress, depressive rumination appears particularly detrimental due to its strong association with hopelessness, avoidance, and prolonged negative affect.

In the context of bereavement, rumination may initially emerge as an attempt to make sense of the loss; however, when persistent, it may hinder adaptive grief processing and contribute to prolonged suffering. Despite growing interest in cognitive processes following bereavement, few studies have specifically examined the role of rumination in perinatal loss (Eisma and Stroebe 2017), and none of them have differentiated between depressive and angry rumination.

The present study aims to address this gap by examining the association between rumination patterns and psychological outcomes following perinatal loss. Specifically, we investigated whether depressive and angry rumination is associated with (1) perinatal grief intensity, (2) depressive and anxiety symptoms, and (3) post-traumatic stress symptoms. We further explored whether combinations of rumination types are associated with greater psychological distress.

## 2 Methods

### 2.1 Study design and participants

The CARING study (Cognitive Analysis and Rumination INvestigation in perinatal Grief) adopted a cross-sectional design. Participants were recruited between December 2023 and May 2024 through online channels of the Foundation. Eligibility criteria included being at least 18 years old and having experienced perinatal loss within the previous five years. The survey was voluntary and anonymous, no personal data were recorded, in no way it was possible to identify the single respondents. Informed consent was provided at the start of the survey once participants had read the participant information and met the eligibility criteria. The study was approved by the Ethics Committee (clinical trial number: not applicable).

### 2.2 Measures

#### 2.2.1 Depressive rumination: Ruminative Response Scale (RRS)

Depressive rumination was assessed using the Ruminative Response Scale (RRS) (Nolen-Hoeksema 1991), a 22-item self-report questionnaire measuring the tendency to engage in repetitive, passive, and self-focused thinking in response to depressed mood. Items assess ruminative thoughts focused on symptoms, causes, and consequences of negative affect and are rated on a 4-point Likert scale. The RRS has shown good reliability and validity across clinical and non-clinical populations and is widely used in research on depression and repetitive negative thinking (Treynor et al. 2003).

#### 2.2.2 Angry rumination: Anger Rumination Scale (ARS)

Angry rumination was measured using the Anger Rumination Scale (ARS) (Sukhodolsky et al. 2001), a 13-item self-report measure assessing the tendency to engage in repetitive thoughts related to anger-provoking experiences. The ARS includes dimensions such as angry afterthoughts, thoughts of revenge, angry memories, and understanding of causes. Items are rated on a 4-point Likert scale. The ARS has demonstrated good psychometric properties and has been used to investigate maladaptive anger-related cognitive processes.

#### 2.2.3 Perinatal grief: Perinatal Grief Scale (PGS)

Perinatal grief was assessed using the Perinatal Grief Scale (PGS) (Potvin et al. 1989), a 33-item self-report questionnaire specifically developed to measure grief responses following perinatal loss. Items are rated on a 5-point Likert scale and yield a total score as well as three subscales: Active Grief, Difficulty Coping, and Despair. The PGS has been extensively used in studies on miscarriage and stillbirth and shows good reliability and construct validity; an Italian language version has been previously validated by our group (Ravaldi et al. 2020).

#### 2.2.4 Psychological distress: Symptom Checklist-90 (SCL-90)

General psychological distress was assessed using the Symptom Checklist-90 (SCL-90) (Derogatis et al. 1973), a 90-item self-report inventory measuring a broad range of psychological symptoms. Items are rated on a 5-point Likert scale and generate a Global Severity Index (GSI) as well as subscale scores, including Depression and Anxiety, which were used in the present study. The SCL-90 is widely validated and commonly used in clinical and research settings.

#### 2.2.5 Post-traumatic stress symptoms: National Stressful Events Survey PTSD Short Scale (NSESSS)

Post-traumatic stress symptoms were measured using the National Stressful Events Survey PTSD Short Scale (NSESSS) (Kilpatrick et al. 2013), a 9-item self-report measure developed by the American Psychiatric Association to assess PTSD symptom severity. Items are rated on a 5-point Likert scale and cover core PTSD domains, including intrusion, avoidance, negative alterations in cognition and mood, and hyperarousal. The NSESSS has shown good reliability and validity in trauma-exposed populations.

### 2.3 Statistical analysis

Given non-normal distributions, non-parametric analyses were employed. Participants were categorized into four groups based on upper-quartile cut-offs for depressive and angry rumination: none, angry rumination only, depressive rumination only, and combined angry and depressive rumination. Group comparisons were conducted using Kruskal–Wallis tests with Dunn post-hoc analyses. Associations between continuous variables were examined using Spearman’s rho.

## 3 Results

### 3.1 Sample characteristics

The final sample comprised 298 participants (97% women), aged 23–48 years. Losses occurred across a wide range of gestational ages, with no significant association between gestational age at loss and rumination levels. Sociodemographic characteristics, loss-related variables, and psychological support received by participants are reported in **Table 1**.

**Table 1.** Sociodemographic and loss-related characteristics of the sample (N = 298)

| ***Characteristic*** | ***n (%)*** |
| --- | --- |
| ***Educational level*** |  |
| *Lower secondary school* | *12 (4.0)* |
| *Upper secondary school* | *91 (30.4)* |
| *Bachelor's degree* | *58 (19.4)* |
| *Master's degree* | *79 (26.4)* |
| *Postgraduate degree* | *58 (19.4)* |
| ***Age at time of loss (years)*** |  |
| *<20* | *4 (1.4)* |
| *21-25* | *37 (12.5)* |
| *26-30* | *111 (37.6)* |
| *31-35* | *112 (38.0)* |
| *36-40* | *29 (9.8)* |
| *41-45* | *2 (0.7)* |
| ***Gestational age at loss (weeks)*** |  |
| *6-19 weeks* | *138 (46.2)* |
| *20-24 weeks* | *60 (20.1)* |
| *25–34 weeks* | *50 (16.7)* |
| *35–42 weeks* | *50 (16.7)* |
| ***Psychological support*** |  |
| *Individual psychotherapy* | *107 (35.9)* |
| *Couple psychotherapy* | *33 (11.0)* |
| *Group psychotherapy* | *8 (2.7)* |
| *Online peer support group* | *25 (8.4)* |
| *In-person peer support group* | *4 (1.3)* |
| *Psychiatric care* | *5 (1.7)* |
| *Counseling* | *5 (1.7)* |
| *Other forms of support* | *11 (3.7)* |

### 3.2 Distribution of rumination profiles

Participants were classified into four rumination profiles based on upper-quartile cut-offs for the Anger Rumination Scale (ARS = 35) and the Ruminative Response Scale (RRS = 53). As illustrated in **Figure 1**, the majority of participants fell into the None group (67.1%, *n* = 200), characterised by low levels of both angry and depressive rumination. The Anger group, defined by elevated angry rumination only, accounted for 8.7% (*n* = 26) of the sample, while the Depression group, characterised by elevated depressive rumination only, comprised 10.7% (*n* = 32). Finally, 13.4% (*n* = 40) of participants exhibited elevated levels of both angry and depressive rumination (Anger & Depression group). Overall, although most participants reported low levels of rumination, a substantial proportion showed elevated angry, depressive, or combined rumination, allowing for meaningful comparisons across distinct cognitive profiles.

**Figure 1.**
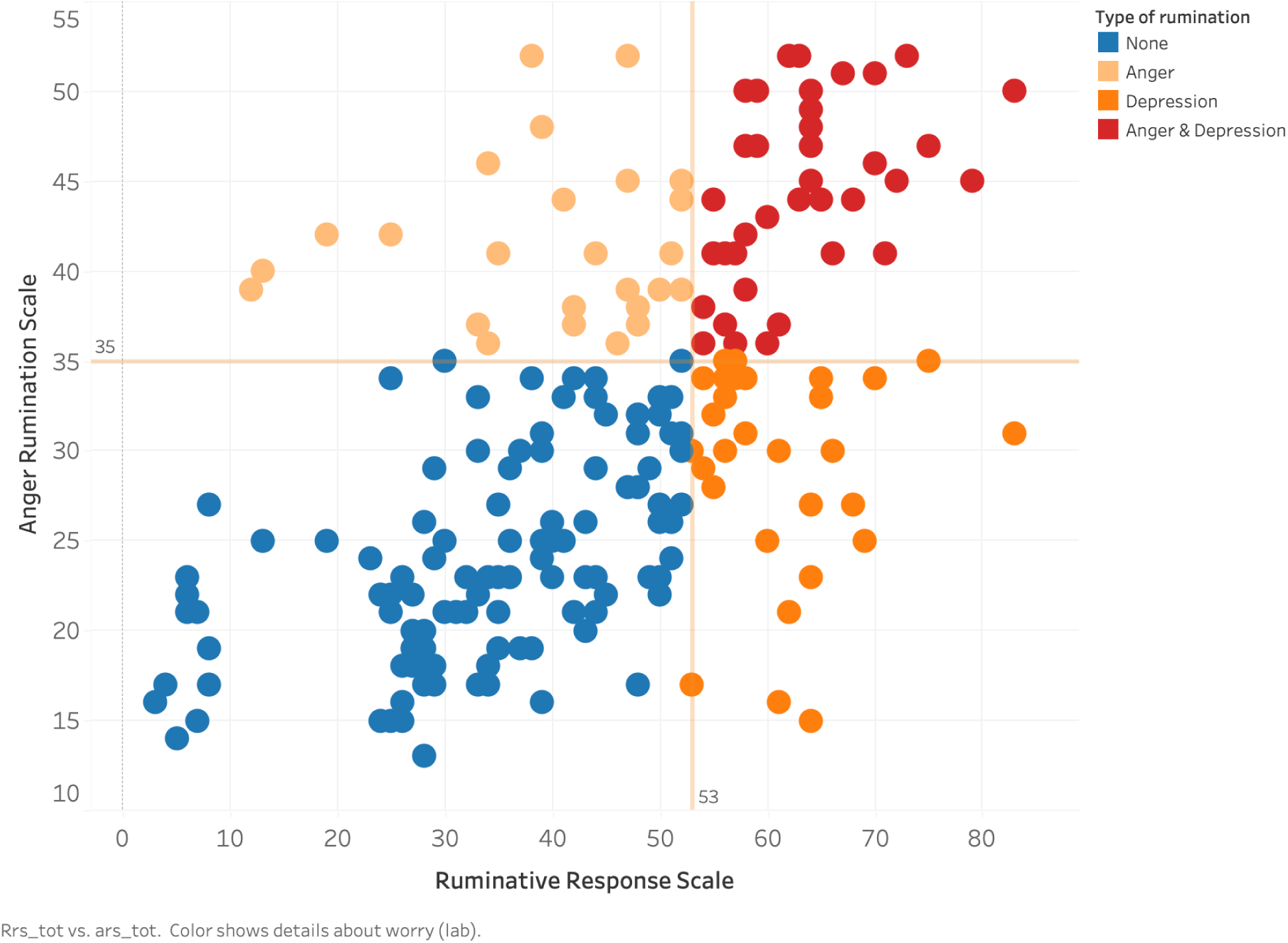
Distribution of participants according to Anger Rumination Scale (ARS) and Ruminative Response Scale (RRS) scores. The scatterplot displays individual participants’ scores on the ARS (y-axis) and RRS (x-axis). Vertical and horizontal lines indicate upper-quartile cut-offs (RRS = 53; ARS = 35), used to classify participants into four rumination profiles: **None**, **Anger**, **Depression**, and **Anger & Depression**. Colours denote rumination profiles.

### 3.3 PTSD severity and rumination

PTSD symptoms, classified into severity categories (low, medium, high, extreme), showed a significantly different distribution across rumination profiles (**Figure 2**). Participants who did not fall within the highest quartile of rumination predominantly reported low or moderate PTSD symptoms, with 44.9% classified as low and 24.4% as medium, while only 10.9% reported extreme symptoms, indicating overall lower levels of post-traumatic distress. In the group characterised by elevated angry rumination, symptom distribution was more evenly spread but shifted towards higher severity levels compared with the None group, with 19.2% in the low category, 34.6% in medium, 23.1% in high, and 23.1% in extreme, suggesting a moderate increase in PTSD severity. Participants with elevated depressive rumination showed a marked concentration of symptoms in the most severe categories: 50.0% were classified as extreme, while only 3.1% fell into the low category, indicating a strong association between depressive rumination and high levels of post-traumatic distress. Finally, the combined angry and depressive rumination profile was associated with the most severe clinical presentation, with 57.5% of participants classified in the extreme category and none in the low category, representing the highest level of PTSD symptom severity among the four groups. Pearson’s chi-square test confirmed that these differences were statistically significant, indicating a robust association between rumination profile and PTSD symptom severity (χ² = 72.86, p < 0.0001).

**Figure 2.**
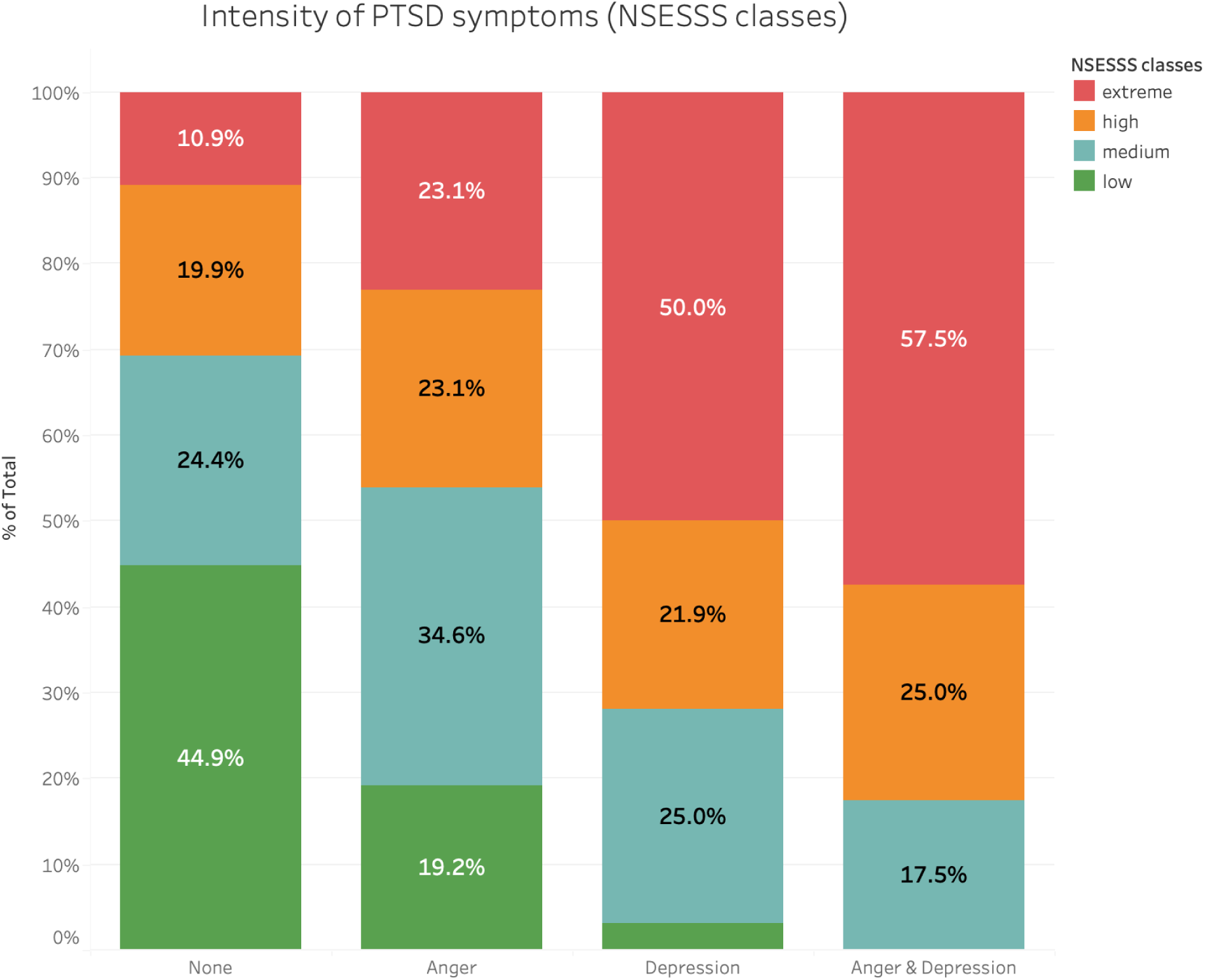
Distribution of PTSD symptom severity (NSESSS classes) across rumination profiles. Stacked bar chart showing the percentage distribution of PTSD severity categories (low, medium, high, extreme) within each rumination group (None, Anger, Depression, Anger & Depression). PTSD severity differed significantly across rumination profiles (χ² = 72.86, p < 0.0001), with depressive and combined rumination associated with a higher proportion of extreme symptoms.

### 3.4 Rumination and psychological distress

Higher levels of rumination were associated with a marked and progressive increase in psychological distress following perinatal loss. As reported in **Table 2**, median scores for perinatal grief, general psychological distress, and post-traumatic stress symptoms increased monotonically across rumination profiles, from participants without rumination to those characterised by angry rumination, depressive rumination, and combined angry and depressive rumination. Differences across groups were statistically significant for all outcomes (Kruskal–Wallis tests: PGS total χ²(3)=79.4; PGS Difficulty Coping χ²(3)=90.9; SCL-90 Depression χ²(3)=61.6; SCL-90 Anxiety χ²(3)=42.7; NSESSS χ²(3)=62.7; all *p*<.001). For total perinatal grief, median PGS scores increased from 89.0 [74.0–103.0] in the no-rumination group to 99.5 [86.0–110.0] in the anger group, 110.5 [98.0–122.0] in the depressive rumination group, and 117.0 [105.0–128.0] among participants with combined rumination (**Online Resource 1**). Post-hoc Dunn tests indicated that both depressive rumination and combined rumination differed significantly from the no-rumination group across all grief outcomes (all *p*<.001), whereas differences between anger-only and depressive rumination were smaller and not consistently significant after adjustment, suggesting a qualitatively stronger association for depressive rumination.

**Table 2.** Median values of perinatal grief, psychological distress, and post-traumatic stress symptoms across rumination profiles.

| **Outcome** | **None<sup>a</sup>**<br>**(n = 200)** | **Anger (A)<sup>a</sup>**<br>**(n = 26)** | **Depression (D)<sup>a</sup>**<br>**(n = 32)** | **A + D<sup>a</sup>**<br>**(n = 40)** | **p<sup>\*</sup>** |
| --- | --- | --- | --- | --- | --- |
| **PGS - Total** | 89.0 [74.0–103.0] | 99.5 [86.0–110.0] | 110.5 [98.0–122.0] | 117.0 [105.0–128.0] | < 0.001 |
| **PGS - Difficulty Coping** | 28.0 [22.0–33.0] | 31.5 [26.0–36.0] | 35.5 [31.0–39.0] | 37.0 [33.0–41.0] | < 0.001 |
| **SCL-90 Depression** | 2.15 [1.70–2.60] | 2.45 [2.00–2.85] | 3.15 [2.70–3.60] | 3.50 [3.00–3.90] | < 0.001 |
| **SCL-90 Anxiety** | 1.80 [1.40–2.20] | 2.05 [1.60–2.50] | 2.60 [2.10–3.05] | 2.95 [2.50–3.40] | < 0.001 |
| **NSESSS** | 14.0 [11.0–18.0] | 17.0 [14.0–21.0] | 22.5 [18.0–27.0] | 24.5 [20.0–29.0] | < 0.001 |

<sup>a</sup>Values are reported as median [25th–75th percentile].
\*p values refer to Kruskal–Wallis tests across rumination profiles.

A similar graded pattern was observed for post-traumatic stress symptoms. Median NSESSS scores rose from 14.0 [11.0–18.0] in participants without rumination to 17.0 [14.0–21.0] in those with angry rumination, 22.5 [18.0–27.0] in those with depressive rumination, and 24.5 [20.0–29.0] in the combined group. Categorical analyses further showed that participants with depressive or combined rumination were disproportionately represented in the high and extreme NSESSS severity classes (χ²(9)=72.9, *p*<.001), whereas individuals without rumination were predominantly classified in the low and medium severity ranges (**Online Resource 2**).

General psychological distress followed the same graded pattern across rumination profiles. Median SCL-90 Depression scores increased from 2.15 [1.70–2.60] in the no-rumination group to 3.50 [3.00–3.90] in the combined rumination group, with analogous increases observed for SCL-90 Anxiety (from 1.80 [1.40–2.20] to 2.95 [2.50–3.40]; **Table 2**). Visual inspection of symptom distributions across rumination profiles is provided in **Online Resource 3**.

### 3.5 Gestational age and rumination

Correlation analyses revealed no significant associations between gestational age at loss and rumination levels, either for angry or depressive rumination. Across the entire sample, correlation coefficients were close to zero (ARS: r = −0.01, p = 0.89; RRS: r = −0.03, p = 0.70), indicating the absence of a linear relationship between gestational age and ruminative thinking. The lack of significant associations was confirmed when analyses were conducted separately for different types of perinatal loss. No meaningful correlations emerged within the miscarriage, termination for fetal anomaly, or intrauterine death groups for either form of rumination. Overall, these findings suggest that gestational age at the time of loss is not associated with levels of angry or depressive rumination, regardless of the type of perinatal loss. Correlation graphs are reported in **Online Resource 4**.

## 4 Discussion

The present study investigated the role of rumination as a cognitive vulnerability factor in psychological adjustment following perinatal loss. Overall, the findings of the CARING study indicate that rumination, particularly depressive rumination and the combined rumination are strongly associated with greater grief severity, higher levels of depression and anxiety, and more severe post-traumatic stress symptoms. These results support and extend existing literature on rumination in bereavement by demonstrating its relevance in the specific and understudied context of perinatal loss.

Consistent with transdiagnostic models of psychopathology, depressive rumination emerged as the most robust and pervasive cognitive correlate of distress across all psychological outcomes examined. Depressive rumination showed stronger associations than angry rumination with perinatal grief intensity, depressive and anxiety symptoms, and PTSD severity. Beyond core affective symptoms, higher levels of rumination were also associated with a broad range of psychological complaints, including somatic symptoms, obsessive–compulsive features, sleep disturbances, and interpersonal sensitivity. This pattern aligns with extensive evidence indicating that depressive rumination, characterized by repetitive, self-focused, and past-oriented thinking, plays a central role in the onset and maintenance of emotional disorders by sustaining negative affect, impairing problem-solving, and reinforcing maladaptive self-appraisals (Nolen-Hoeksema 1991; Watkins and Brown 2002; Watkins 2008).

Previous studies have shown that rumination following bereavement is associated with loss avoidance and poorer psychological adjustment (Eisma et al. 2014). In this context, rumination has been conceptualized as a cognitive process that may initially serve an attempt at meaning-making but becomes maladaptive when it remains abstract, repetitive, and emotionally saturated (Eisma and Stroebe 2017). The present findings suggest that this mechanism is particularly salient in perinatal loss, where parents are confronted not only with the death of a child but also with the collapse of anticipated identities, relational roles, and future narratives. Persistent depressive rumination may therefore interfere with adaptive grief processing by maintaining attention on loss-related discrepancies and counterfactual thoughts, preventing integration of the loss into autobiographical memory.

The progressive increase in grief severity, psychological distress, and post-traumatic stress symptoms across rumination profiles indicates that higher levels of rumination are associated with greater psychological burden following perinatal loss. Importantly, this pattern suggests a continuum rather than a dichotomous distinction between adaptive and maladaptive cognitive responses, with increasing levels of ruminative engagement corresponding to progressively greater psychological burden.

Participants presenting both depressive and angry rumination exhibited the highest levels of grief severity, psychological distress, and PTSD symptoms. These findings suggest that co-occurring ruminative processes may interact synergistically, amplifying emotional suffering. While angry rumination alone was associated with increased distress compared to the absence of rumination, its impact was consistently weaker than that of depressive rumination, in line with previous research indicating that anger-focused repetitive thinking may be more situational and externally oriented, and therefore less persistent over time (Bushman et al. 2001; Watkins 2008).

The strong association between rumination and post-traumatic stress symptoms represents a particularly relevant finding. Participants characterized by depressive or combined rumination tended to cluster in the higher PTSD severity categories, whereas those with angry rumination alone or no elevated rumination were more commonly represented in lower severity levels.

This result is consistent with cognitive models of PTSD, which identify repetitive negative thinking as a key mechanism sustaining intrusive memories, hyperarousal, and avoidance (Ehlers and Clark 2000). In perinatal loss, ruminative thinking may repeatedly reactivate traumatic representations of the loss event while simultaneously preventing emotional processing, thereby increasing vulnerability to persistent post-traumatic stress.

Importantly, rumination was not associated with gestational age at the time of loss. This finding supports prior evidence indicating that the psychological impact of perinatal loss is more strongly related to emotional attachment and meaning attribution than to biological timing or medical classification of the loss (Kersting and Wagner 2012). From a clinical perspective, this result underscores the importance of avoiding assumptions about the severity of parental distress based solely on gestational age and instead focusing on cognitive-emotional processes that may influence adjustment trajectories.

Contrary to expectations, no significant differences in rumination levels were observed between participants who reported having received psychological support and those who had not. This finding should be interpreted cautiously. The study did not assess the type, duration, timing, or theoretical orientation of psychological interventions, limiting conclusions regarding treatment effectiveness. It is possible that many interventions were not specifically designed to target ruminative processes, which are known to be particularly resistant to change and may require structured, process-focused approaches such as metacognitive therapy or rumination-focused cognitive-behavioral interventions (Watkins et al. 2007; Wells 2009). Rather than suggesting scarce efficacy of psychological support per se, this result highlights the need for interventions that explicitly address maladaptive rumination in the aftermath of perinatal loss.

Taken together, the findings of this study support the conceptualization of depressive rumination as a transdiagnostic cognitive mechanism contributing to persistent distress following perinatal bereavement. By extending rumination research to the perinatal domain, the present study contributes to a more process-oriented understanding of grief-related psychopathology and identifies rumination as a clinically relevant target for assessment and intervention.

### 4.1 Clinical implications

The identification of rumination, particularly depressive rumination, as a central vulnerability factor has important clinical implications. Early screening for ruminative tendencies in parents following perinatal loss may help identify individuals at increased risk for prolonged grief, depression, and PTSD. Interventions that explicitly aim to reduce ruminative thinking, promote concrete and adaptive coping strategies, and facilitate emotional processing may be particularly beneficial in this population. Integrating rumination-focused components into perinatal bereavement care may therefore improve psychological outcomes and reduce long-term distress.

Consistent with this process-oriented perspective, psychotherapeutic interventions specifically targeting ruminative processes appear particularly appropriate. Metacognitive Therapy (MCT) directly addresses dysfunctional metacognitive beliefs that sustain repetitive negative thinking, reducing the perception of rumination as a useful or necessary strategy for coping with loss (Wells and Matthews 1994; Papageorgiou and Wells 2003; Wells 2009). Acceptance and Commitment Therapy (ACT), by contrast, focuses on altering individuals’ relationships with painful thoughts and emotions by reducing cognitive fusion and experiential avoidance, while promoting values-oriented adaptation even in the presence of suffering (Hayes et al. 2012; Levin et al. 2024)

In addition, Eye Movement Desensitization and Reprocessing (EMDR) may be especially indicated when ruminative processes are maintained by unresolved traumatic memories and intrusive imagery related to the loss, facilitating adaptive reprocessing of the event and potentially reducing cognitive perseveration (World Health Organization 2013).

Overall, the findings of the present study suggest that the severity of psychological distress following perinatal loss does not primarily depend on gestational age at the time of loss, but rather may be amplified by cognitive and emotional processes, particularly rumination. This observation reinforces the importance of a process-oriented clinical approach capable of recognising subjective suffering beyond temporal or medical criteria, and of targeting mechanisms that play a substantial role in the maintenance of emotional distress.

## 5 Strengths and limitations

This study should be interpreted in light of several strengths and limitations. Among its main strengths is the focus on rumination as a transdiagnostic cognitive process in the context of perinatal loss, an area that remains relatively understudied despite its clinical relevance. By distinguishing between depressive rumination, angry rumination, and their co-occurrence, the study offers a nuanced and process-oriented understanding of cognitive vulnerability beyond categorical diagnostic frameworks. In addition, the use of multiple validated measures allowed for a comprehensive assessment of grief, general psychological distress, and post-traumatic stress symptoms, capturing both outcome severity and symptom profiles across domains. The relatively large sample size for this population and the inclusion of losses across a wide range of gestational ages further strengthen the robustness and generalizability of the findings within perinatal bereavement contexts. Importantly, the consistency of results across continuous, categorical, and distributional analyses supports the stability of the observed associations.

Several limitations should also be acknowledged. The cross-sectional design precludes causal inferences regarding the directionality of associations between rumination and psychological outcomes. The sample was predominantly female, limiting generalizability to fathers and other partners, whose experiences of rumination and grief may differ. Moreover, psychological support was assessed in a broad and non-specific manner, without information on timing, duration, or therapeutic orientation, preventing conclusions about the effectiveness of different interventions. Future studies should adopt longitudinal designs to examine the temporal dynamics of rumination following perinatal loss and to evaluate whether changes in ruminative thinking predict recovery trajectories. Further research is also needed to test the effectiveness of targeted, rumination-focused interventions in reducing distress among bereaved parents.

## 6 Conclusions

The present study highlights rumination as a central cognitive process associated with persistent psychological distress following perinatal loss. Depressive rumination, alone or in combination with angry rumination, emerged as the most strongly associated with grief severity, depressive and anxiety symptoms, and post-traumatic stress, underscoring its relevance as a transdiagnostic vulnerability factor in perinatal bereavement. By demonstrating that psychological suffering is more closely related to ruminative processes than to gestational age at the time of loss, these findings challenge medically or temporally driven assumptions about grief severity and support a more individualised, process-oriented understanding of parental adjustment after loss. From this perspective, early identification of ruminative tendencies may help to identify parents at increased risk for prolonged distress, independently of the clinical characteristics of the loss.

Interventions aimed at reducing maladaptive rumination and promoting adaptive modes of emotional processing may therefore play a crucial role in perinatal mental health care. Integrating rumination-focused assessment and treatment components into bereavement support pathways could contribute to more targeted, responsive, and effective interventions for parents experiencing perinatal loss.

## Data Availability

All data produced in the present study are available upon reasonable request to the authors

## 8 Author contributions

**C.R.**: Conceptualization, Methodology, Writing-Original Draft; **L.M.:** Conceptualization, Methodology, Writing-Review & Editing; **G.R.:** Conceptualization, Methodology, Writing-Review & Editing; **C.O.:** Conceptualization, Methodology, Writing-Review & Editing; **I.N.:** Conceptualization, Methodology, Writing-Review & Editing; **E.C.:** Conceptualization, Methodology, Writing-Review & Editing; **A.V.:** Conceptualization, Methodology, Formal Analysis, Data Curation, Writing-Review & Editing, Supervision.

## Acknowledgments

CiaoLapo Foundation for Healthy Pregnancy and Perinatal Loss Support provided infrastructure for the realisation of the study (documents, questionnaires, material, software, web platforms, open access, etc.).

## Declarations

### Ethics approval

The study was approved by the Ethics Committee of the University of Florence (protocol no. 0035322, February 14, 2024).

