## Supplementary files for "Rumination as a cognitive vulnerability factor in perinatal bereavement: evidence from the CARING study"

Supplementary Materials

#### Supplementary Material 1. Additional statistical analyses

##### Correlation matrices

To further explore the relationships between rumination and psychological outcomes, correlation analyses were conducted between continuous scores on the Anger Rumination Scale (ARS), the Ruminative Response Scale (RRS), the Perinatal Grief Scale (PGS) and its subscales, the Symptom Checklist-90 (SCL-90), and the National Stressful Events Survey PTSD Short Scale (NSESSS).

Given the non-normal distribution of several variables, Spearman's rank-order correlation coefficients (ρ) were used. The results indicate moderate to strong positive correlations between rumination—particularly depressive rumination—and all psychological outcome measures, including perinatal grief severity, depressive and anxiety symptoms, and post-traumatic stress symptoms. Correlations involving depressive rumination were consistently stronger than those involving angry rumination.

The full correlation matrix is reported in Table S1.

#### Supplementary Material 2. Additional group comparisons

Additional non-parametric group comparisons were conducted to further characterize differences across rumination profiles (None, Anger, Depression, Anger & Depression).

##### Psychological distress (SCL-90)

Participants characterized by depressive rumination or combined angry and depressive rumination showed significantly higher scores on the SCL-90 Depression and Anxiety subscales compared to participants without elevated rumination. While angry rumination alone was associated with increased depressive symptoms relative to the absence of rumination, its impact was consistently weaker than that observed for depressive rumination.

Detailed results of Kruskal–Wallis tests and Dunn post-hoc comparisons are reported in Table S2.

#### Supplementary Material 3. Gestational age and rumination

Exploratory analyses examined the association between gestational age at loss and levels of depressive and angry rumination. No significant correlations were observed between gestational age and ARS or RRS scores, either in the overall sample or within loss subgroups. These findings suggest that rumination levels are independent of the biological timing of the loss.

Corresponding correlation coefficients are reported in Table S3.

#### Supplementary Material 4. Psychological support and rumination

Participants were also compared based on whether they reported having received psychological support following the loss. No significant differences in ARS or RRS scores were observed between participants who reported receiving psychological support and those who did not. Given the lack of information regarding the type, duration, or timing of interventions, these findings should be interpreted cautiously.

Group comparison statistics are reported in Table S4.

#### Supplementary Tables

- Table S1. Spearman correlation matrix between ARS, RRS, PGS (total and subscales), SCL-90 (GSI, Depression, Anxiety), and NSESSS

• Table S2. Kruskal–Wallis and Dunn post-hoc comparisons for SCL-90 Depression and Anxiety across rumination profiles

• Table S3. Correlations between gestational age at loss and rumination scores

• Table S4. Comparison of rumination scores by psychological support status

### Supplementary Tables

#### Table S1. Spearman correlation matrix between rumination scales and psychological outcomes

| | 1 | 2 | 3 | 4 | 5 | 6 | 7 | 8 | 9 | 10 |
|---|---|---|---|---|---|---|---|---|---|---|
| 1. ARS | 1.00 | | | | | | | | | |
| 2. RRS | 0.61*** | 1.00 | | | | | | | | |
| 3. PGS Total | 0.50*** | 0.61*** | 1.00 | | | | | | | |
| 4. PGS Active Grief | 0.41*** | 0.51*** | 0.89*** | 1.00 | | | | | | |
| 5. PGS Difficulty Coping | 0.53*** | 0.64*** | 0.92*** | 0.72*** | 1.00 | | | | | |
| 6. PGS Despair | 0.46*** | 0.56*** | 0.95*** | 0.78*** | 0.84*** | 1.00 | | | | |
| 7. SCL-90 GSI | 0.61*** | 0.66*** | 0.69*** | 0.57*** | 0.72*** | 0.64*** | 1.00 | | | |
| 8. SCL-90 Depression | 0.57*** | 0.65*** | 0.70*** | 0.59*** | 0.75*** | 0.62*** | 0.92*** | 1.00 | | |
| 9. SCL-90 Anxiety | 0.52*** | 0.55*** | 0.57*** | 0.47*** | 0.60*** | 0.53*** | 0.92*** | 0.82*** | 1.00 | |
| 10. NSESSS | 0.52*** | 0.65*** | 0.65*** | 0.55*** | 0.63*** | 0.63*** | 0.73*** | 0.67*** | 0.68*** | 1.00 |

Values are Spearman rank-order correlation coefficients (ρ). *** $p<0.001$, ** $p<0.01$, * $p<0.05$. Coefficients computed on pairwise-complete observations. ARS, Anger Rumination Scale; RRS, Ruminative Response Scale; PGS, Perinatal Grief Scale; SCL-90, Symptom Checklist-90; GSI, Global Severity Index; NSESSS, National Stressful Events Survey PTSD Short Scale.

#### Table S2. Kruskal–Wallis tests and Dunn post-hoc comparisons for SCL-90 Depression and Anxiety across rumination profiles

| Outcome | None (n=97) | Anger (n=20) | Depression (n=27) | A+D (n=36) | H (df=3) | p |
|---|---|---|---|---|---|---|
| SCL-90 Depression | 2.15 [1.69–2.77] | 2.73 [2.23–3.54] | 3.15 [2.81–3.46] | 3.50 [3.00–4.08] | 61.67 | <0.001 |
| SCL-90 Anxiety | 1.80 [1.40–2.30] | 2.20 [1.95–2.62] | 2.60 [2.15–3.00] | 2.95 [2.20–3.60] | 42.81 | <0.001 |

**Dunn post-hoc (Bonferroni-adjusted p):**
*SCL-90 Depression* — None vs Anger: z=-2.66, p=0.047; None vs Depression: z=-4.82, p=<0.001; None vs A+D: z=-7.15, p=<0.001; Anger vs Depression: z=-1.34, p=1.000; Anger vs A+D: z=-2.66, p=0.047; Depression vs A+D: z=-1.36, p=1.000
*SCL-90 Anxiety* — None vs Anger: z=-1.97, p=0.291; None vs Depression: z=-4.05, p=<0.001; None vs A+D: z=-5.96, p=<0.001; Anger vs Depression: z=-1.34, p=1.000; Anger vs A+D: z=-2.43, p=0.090; Depression vs A+D: z=-1.11, p=1.000
Values are median [25th–75th percentile]. H, Kruskal–Wallis statistic.

#### Table S3. Correlations between gestational age at loss and rumination scores

| Subgroup | ARS ρ (p, n) | RRS ρ (p, n) |
|---|---|---|
| Overall | -0.01 (p=0.869, n=125) | -0.04 (p=0.626, n=143) |
| Miscarriage (<20 wk) | n/a (n=2) | n/a (n=2) |
| Termination for fetal anomaly | -0.10 (p=0.577, n=31) | 0.02 (p=0.912, n=34) |
| Intrauterine death (≥20 wk) | 0.05 (p=0.672, n=67) | 0.00 (p=0.990, n=77) |

Spearman rank-order correlation coefficients between gestational age at loss (in weeks, recorded for losses ≥20 weeks) and rumination scores. The miscarriage subgroup is not applicable (n/a) because gestational age in weeks was recorded only for losses ≥20 weeks; n differs between scales owing to item-level missingness. ARS, Anger Rumination Scale; RRS, Ruminative Response Scale.

#### Table S4. Rumination scores by psychological support status

| Scale | No support | Support received | p* |
|---|---|---|---|
| ARS | 30.0 [22–39] (n=98) | 30.0 [23–37] (n=116) | 0.857 |
| RRS | 41.5 [30–55] (n=114) | 48.0 [32–57] (n=133) | 0.183 |

Values are median [25th–75th percentile]. *Mann–Whitney U test (two-sided). ARS, Anger Rumination Scale; RRS, Ruminative Response Scale.

### Online Resource (Figures)

**Online Resource 1. Distribution of Perinatal Grief Scale (PGS) scores across rumination profiles.** Boxplots show median values, interquartile ranges, and distribution of scores for Total PGS, Active Grief, Difficulty Coping, and Despair subscales in participants with no elevated rumination, angry rumination, depressive rumination, and combined angry and depressive rumination. Higher levels of rumination, particularly depressive and combined rumination, were associated with progressively higher grief scores across all PGS dimensions.

**Online Resource 2. Post-traumatic stress symptom severity (NSESSS) by rumination profile.** Distribution of post-traumatic stress symptom severity according to rumination profile. Boxplots represent NSESSS total scores across participants with no elevated rumination, angry rumination, depressive rumination, and combined angry and depressive rumination. Participants with depressive and combined rumination showed higher median NSESSS scores and greater dispersion toward the upper range, consistent with increased representation in high and extreme PTSD severity categories.

**Online Resource 3. SCL-90 symptom dimensions by rumination profile.** Distribution of SCL-90 symptom dimensions across rumination profiles. Boxplots depict Global Severity Index (GSI) and individual symptom domains (somatization, obsessive–compulsive, interpersonal sensitivity, depression, anxiety, hostility, phobic anxiety, paranoid ideation, psychoticism, and sleep disturbances) for participants with no elevated rumination, angry rumination, depressive rumination, and combined angry and depressive rumination. Across domains, higher rumination levels, particularly depressive and combined rumination, were associated with higher symptom severity.

**Online Resource 4. Relationship between gestational age at loss and rumination levels.** Scatterplots showing the association between gestational age at loss and (A) angry rumination (Anger Rumination Scale, ARS) and (B) depressive rumination (Ruminative Response Scale, RRS). Colours indicate type of perinatal loss (miscarriage <20 weeks, intrauterine death ≥20 weeks, termination of pregnancy for fetal anomaly). No significant correlations were observed between gestational age and rumination levels.

**Online Resource 1.**

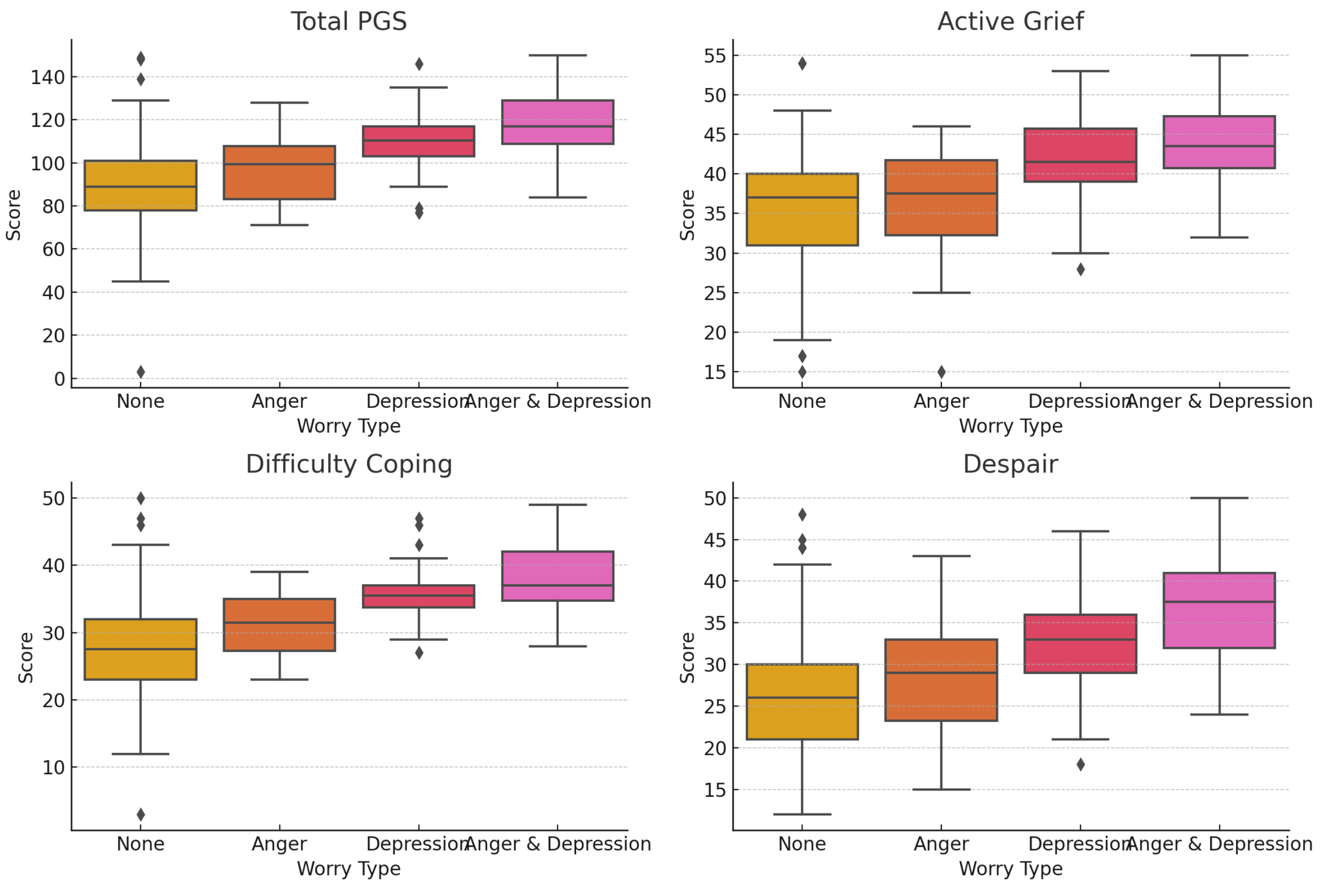

**Online Resource 2.**

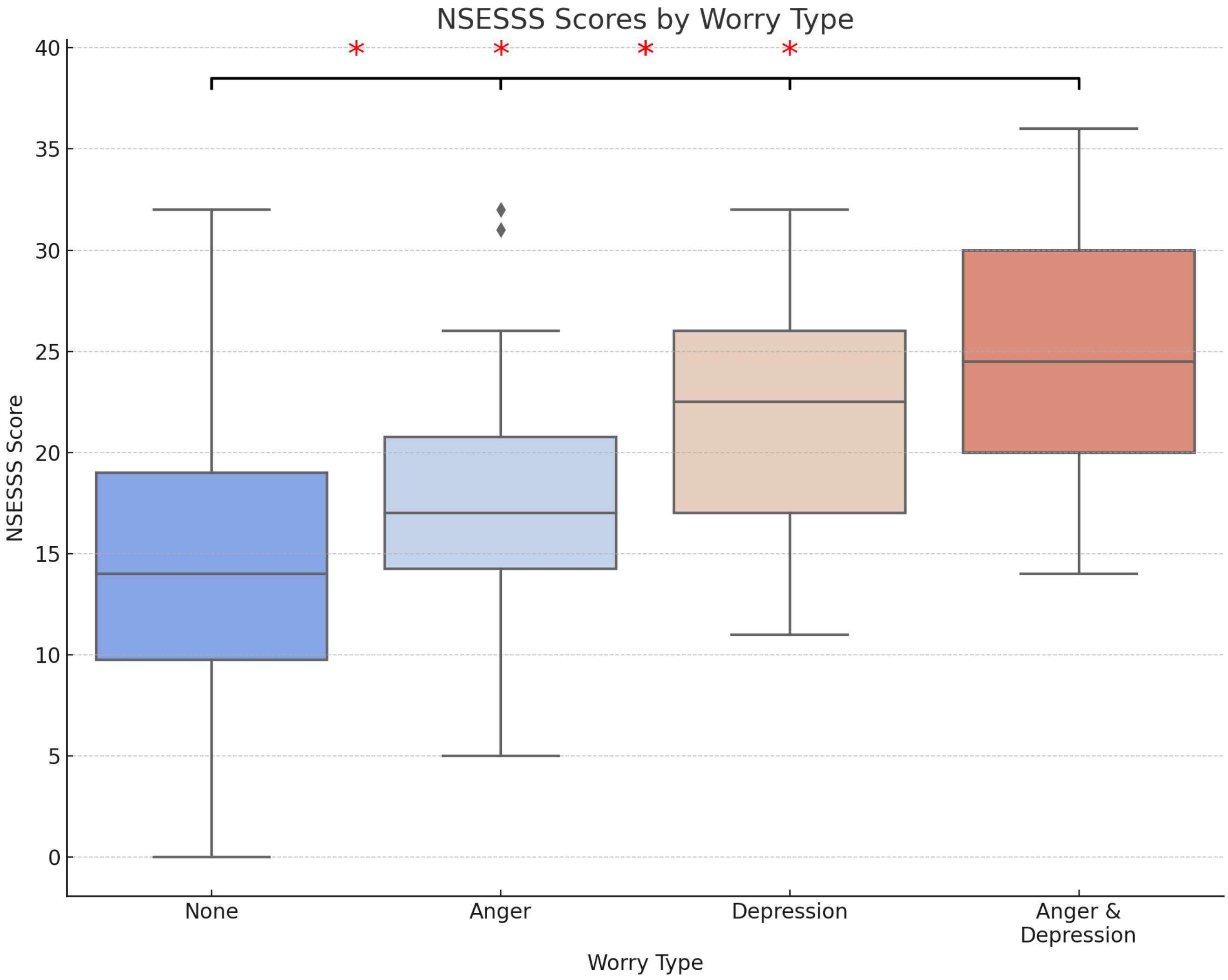

### Online Resource 3.

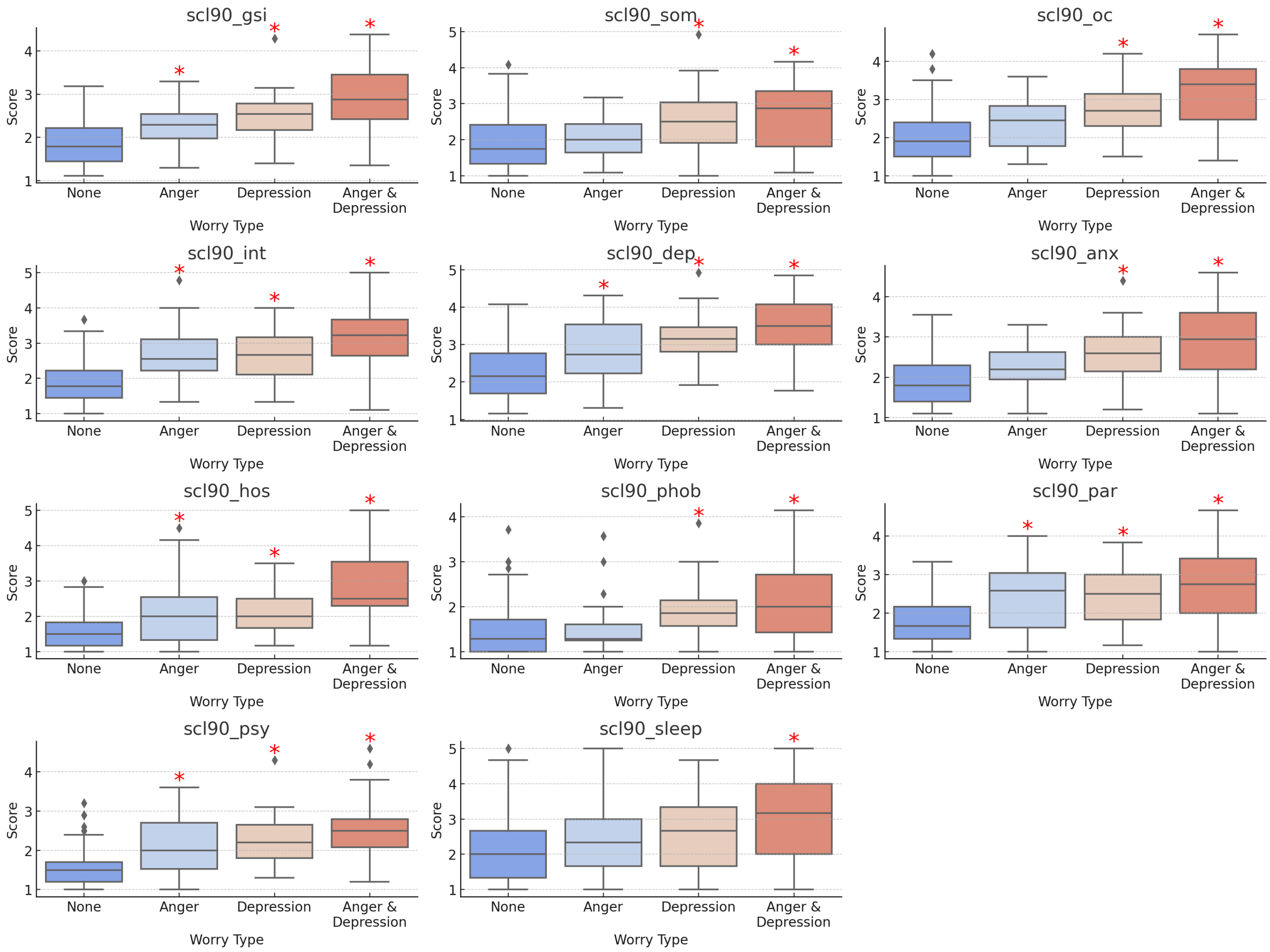

**Online Resource 4.**

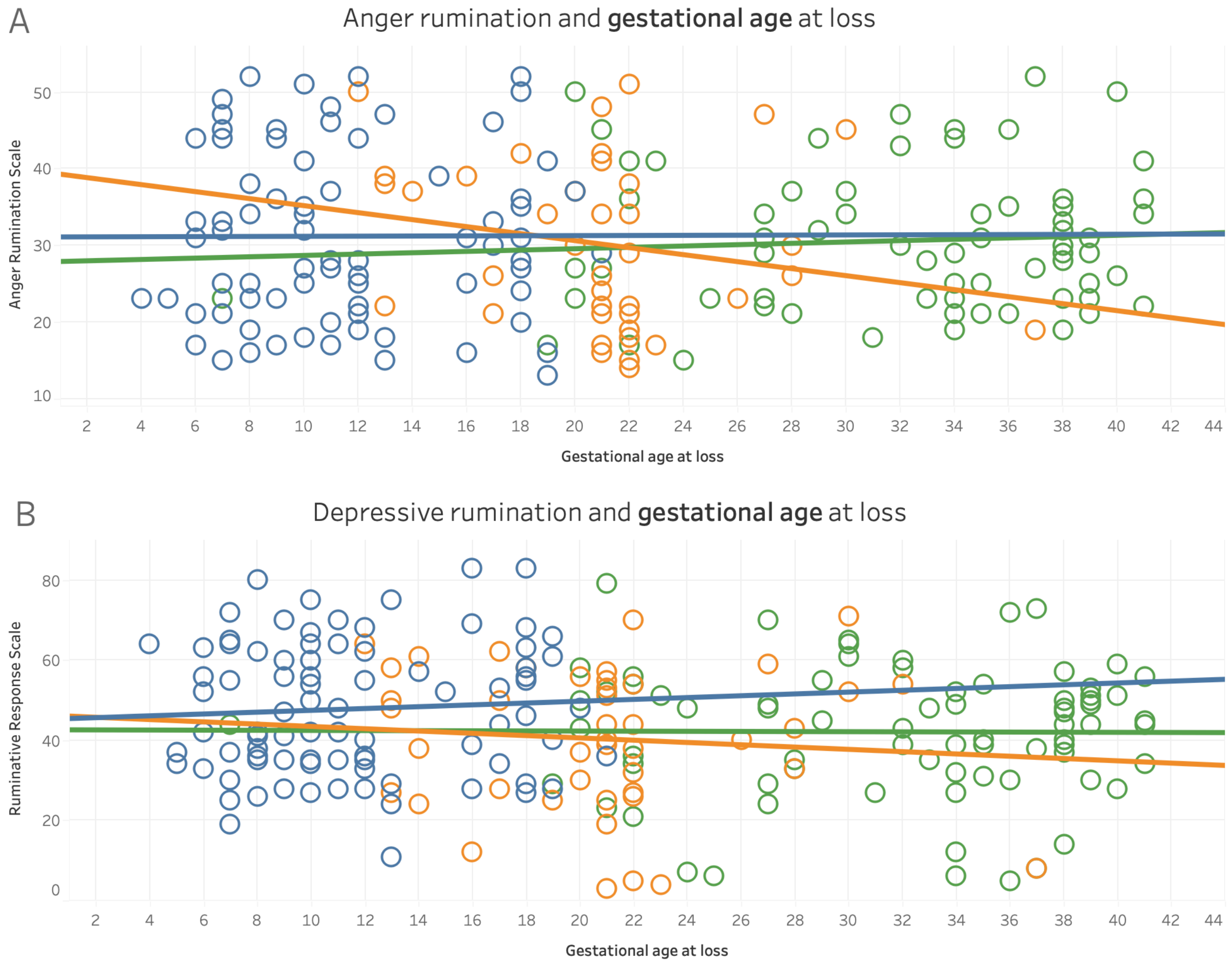
